# Asexual stage synchronicity in symptomatic and asymptomatic falciparum malaria

**DOI:** 10.1101/2022.03.13.22272117

**Authors:** James A Watson, Nicholas J White

## Abstract

Andrade et al have reported that *P. falciparum* parasitised erythrocytes circulate for longer in persistent asymptomatic infections than in symptomatic malaria. This radical suggestion, attributed to in-vivo adaptation by the parasite population to reduced cytoadherence, is based largely on in-vivo transcriptomic data from 24 Malian children: 12 with acute falciparum malaria and 12 with asymptomatic parasitaemia. We show that the reported analysis generated erroneous results because of data formatting issues. We also show that the algorithm used to estimate the average asexual parasite developmental stage (hours post-invasion) from in-vivo transcriptomic data breaks down when applied to asynchronous infections. We argue that comparisons between asymptomatic and symptomatic malaria of asexual parasite developmental stage distributions are confounded by differences in synchronicity and gametocytaemia, and also by selection bias (because schizogony often precipitates clinical presentation). There is no convincing evidence of an adaptive delayed cytoadherence phenotype in chronic *P. falciparum* infections.

## 1 Background

In the study by Andrade *et al* blood samples from children presenting with their first symptomatic malaria infection of the rainy season were compared with samples from children who had asymptomatic infections persisting during the dry season [1]. The authors suggest that *P. falciparum* somehow adapts to chronicity by reducing its endothelial binding, and that splenic clearance is the main mechanism maintaining low parasitaemias during the dry season. Central to this hypothesis are parasite RNA sequencing data from 12 asymptomatic Malian children (dry season) and 12 symptomatic children (wet season). Both groups were selected for intensive study because they had relatively high parasitaemias (the highest densities observed in the wet and dry season cohorts) [1].

## 2 Reproducibility

We downloaded the RNA seq data from the National Center of Biotechnology Information’s Gene Expression Omnibus (GEO, accession number GSE148125) and the *in vitro* reference asexual transcriptome data [2]. We used the R script provided by the authors which implements maximum likelihood estimation (MLE) of the parasite age (hours post invasion: hpi) based on the reference transcriptome published by Bozdech *et al* (code is provided in the accompanying github repository) [2, 3]. We could not replicate the published hpi estimates. The greatest differences were observed for the dry season samples (Figure 1). After contacting the authors, we discovered that their original code had loaded the data in csv format whereby the decimal point was coded using the European convention of a comma instead of a dot. As a result their code had interpreted the RNA seq data as character strings instead of numbers, resulting in semi-arbitrary outputs from the MLE algorithm (Figure 1: correlation between results using the numerical data and the results using the character string data is because the underlying algorithm works on the ranks of the gene expression data, and there is some correlation between the order of the digits within each number and the size of the number^1^).

**Fig. 1.**
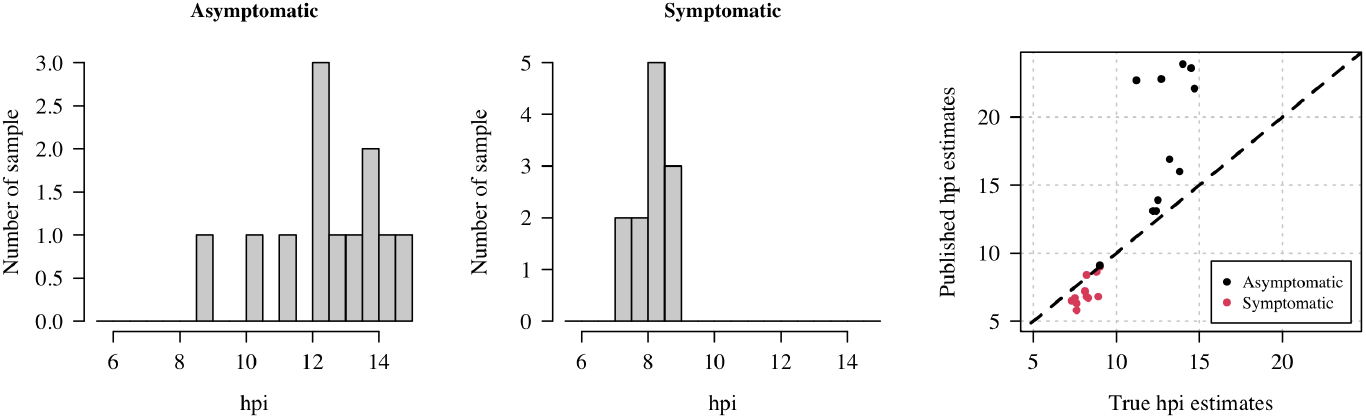
Updated maximum likelihood estimates of the hours post invasion for the 12 asymptomatic infections (left) and the 12 symptomatic infections (middle). A comparison between our updated estimates (x-axis) and the published estimates (y-axis) is shown in the right panel.

Our re-analysis of their RNA seq data is consistent with the reported *ex vivo* experiments. Under the model [3], the parasites in the asymptomatic infections are slightly older than those in the symptomatic infections (Figure 1). The difference in median ages is 4.5 hours, not 10 hours as given in the published analysis. The oldest mean estimated age is ~15 hpi (instead of 24 hpi), consistent with the general view that sequestration starts before parasites mature into large rings (16 hours into the lifecycle). The MLE estimates for the symptomatic infections are tightly clustered around 8 hours.

## 3 Comparison of developmental stages in asymptomatic and symptomatic infections

Parasite developmental stage comparisons between asymptomatic infections and the selected symptomatic infections are inherently confounded by two factors.

First, there is selection bias with respect to developmental stage in symptomatic malaria. Fever in falciparum malaria is precipitated by schizogony, influencing treatment seeking behavior [4]. Patients with uncomplicated falciparum malaria usually present shortly after fever onset. This explains the observation, noted for over a century, that patients with acute falciparum malaria usually present with a predominance of young ring form parasites in their peripheral blood films. The bias to young ring stage parasites in the report by Andrade *et al* is compounded by selection of the 12 patients with the highest parasite densities: as Fairley showed in his seminal studies, peripheral blood parasitaemia in acute falciparum malaria follows an increasing sine-wave pattern driven by synchronous iterations of schizogony and sequestration [5]. This sine-wave pattern has been demonstrated more recently in human challenge studies [6]. If parasitaemia follows a sine-wave pattern driven by the mean developmental age, it follows that selecting patients with the highest peripheral parasitaemia will select for those with highly synchronous infections following schizogony [7].

Second, we do not expect the degree of parasite developmental synchronicity to be the same in asymptomatic and symptomatic malaria. In persistent asymptomatic malaria (by definition in the absence of fever), we expect an asynchronous infection, with little correlation in hpi across infected red cells. In contrast, in uncomplicated malaria, fever synchronises the infection by predominantly inhibiting growth of mature stage parasites [8]. When we compare the distributions of gene expressions in asymptomatic infections versus symptomatic infections, it is clear that the expression profiles in the asymptomatics have greater variance relative to the symptomatics (i.e. a greater variety of genes are expressed) consistent with more asynchronous infections (Figure 2). The sample size in Andrade *et al* is extremely small: a 12 versus 12 comparison. If the asymptomatic infections were substantially asynchronous as might be expected, then this in itself could explain the presence of older circulating parasites.

**Fig. 2.**
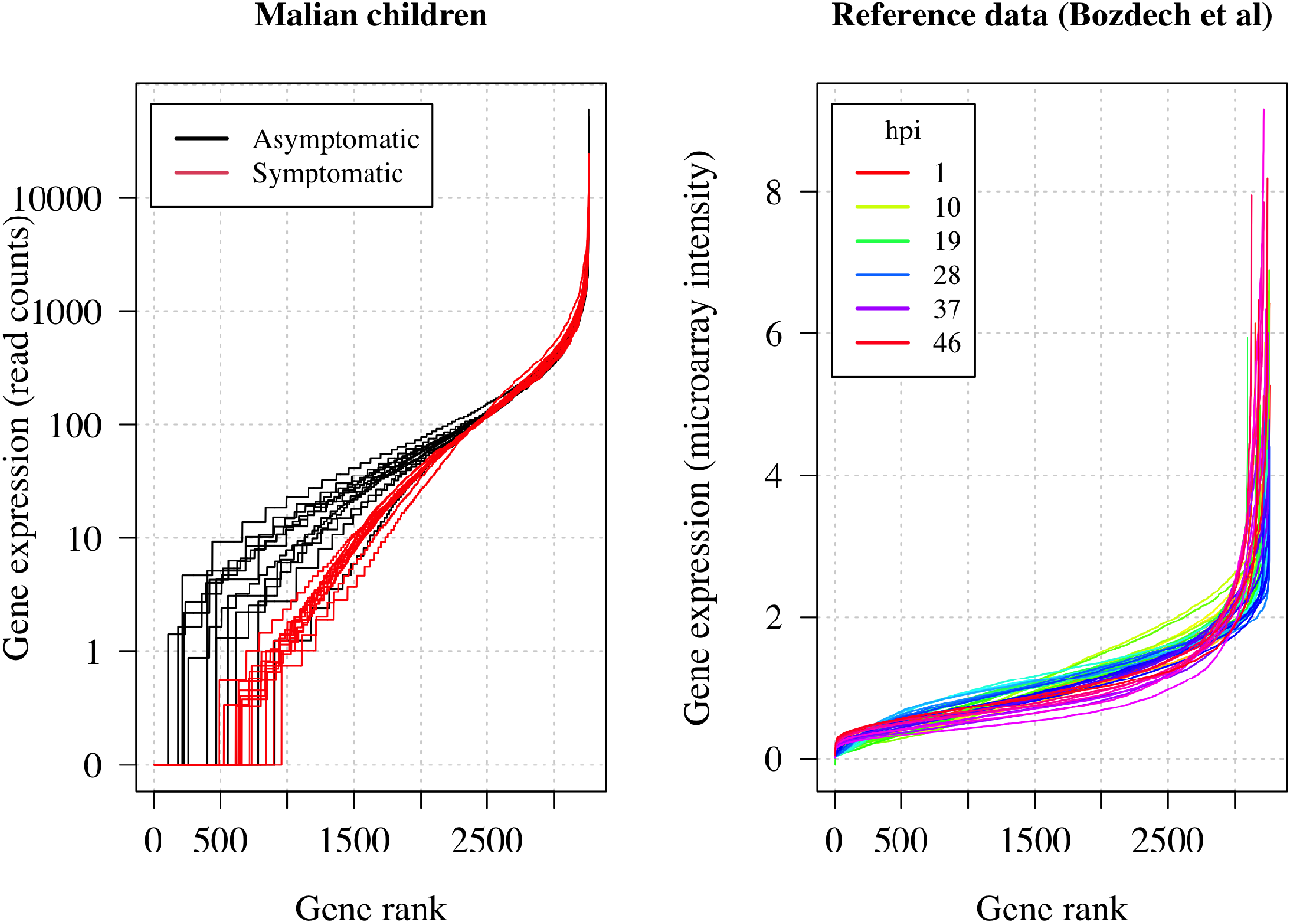
In both panels, we have ranked the genes from least expressed (rank 0) to most expressed, and then plotted the corresponding expression value. For the patient data, the y-axis shows the number of RNA read counts (log scale); for the reference data [2], the y-axis shows the 2-color microarray intensity values. The asymptomatic infections in Malian children have a wider range of genes with non-zero read counts, consistent with a wider range of parasite ages. In the reference data, at all stages we see a highly consistent proportion of genes differentially expressed.

In summary, differences in hpi between asymptomatic and symptomatic malaria could in theory be explained by selection bias and by differences in synchronicity.

## 4 Estimating hpi in asynchronous infections

In addition to systematic confounding, it is important to characterize the performance of the MLE algorithm in asynchronous infections. We show using simulated data that the underlying algorithm for estimating hpi is highly sensitive to the assumption of synchronicity.

We simulated aggregate microarray gene expression data from 10,000 circulating parasites using the published reference genome [2], varying the mean parasite age from 1 to 15 hours and the standard deviation of the parasite age distribution within the host from 1 to 13 hours, under the assumption that parasite ages are normally distributed within the host (modulo 48 hours). The simulations were done with and without sequestration (see supplementary code). Both with and without sequestration in the model we observed pathological behavior of the algorithm for large standard deviation values, i.e. highly asynchronous infections (Figure 3). As we increase asynchrony in the simulated infection (i.e. increase the standard deviation of the age distribution), the hpi estimates converge to fixed time-points, depending on whether we include sequestration and depending on the simulated mean age. When we include sequestration in the simulated data, the estimated hpi converges to 10 hours, irrespective of the true mean age in the simulation. This is unsurprising: the model from Lemieux *et al* does not consider the gene expression data as an aggregate value from multiple parasites with a distribution of ages, nor does it consider sequestration of later stage parasites. It considers the observed *in vivo* data as an unbiased single noisy realization of the reference transcriptome [3]. This assumption may be approximately true in symptomatic malaria, but is unlikely to hold in asymptomatic infections.

**Fig. 3.**
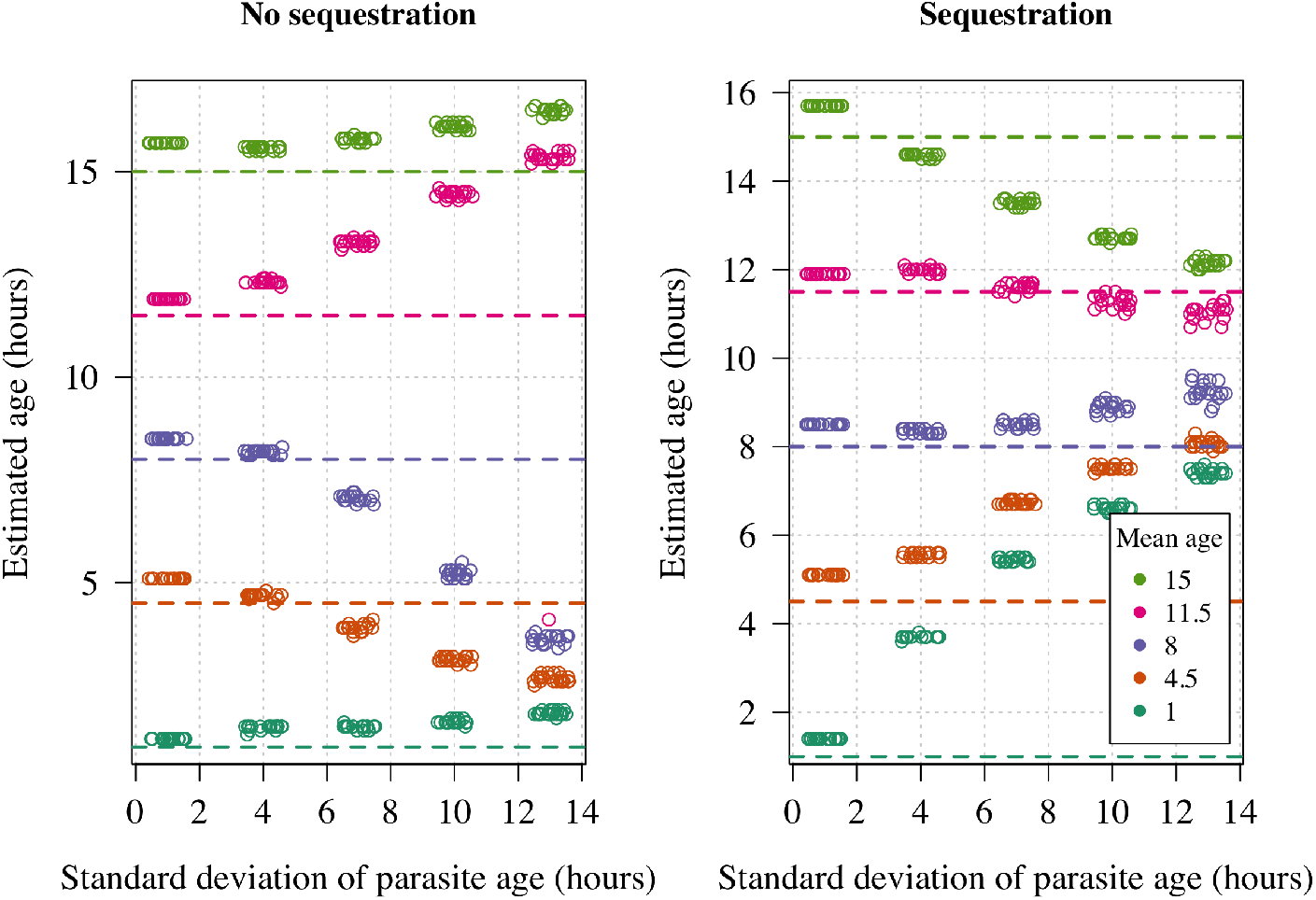
Simulation study to understand the effect of asynchrony on the hours post invasion estimate using the method from Lemieux *et al* [3]. The no sequestration simulation assumed that parasites of all ages are in circulation; the sequestration experiment assumed that infected red cells start to sequester after 16 hours (large rings) at a constant rate, and by 38 hours only 1 in a thousand infected red cells are circulating. The sequestration parameters in the simulation are speculative and serve only to illustrate the sensitivity of the algorithm to input data.

Another aspect to consider is the influence of circulating gametocytes on the estimated hpi. The published method from Lemieux *et al* [3] proposed using a mixture model to correct for possible contamination from gametocyte transcription. However, in the analysis by Andrade *et al* gametocyte gene expression is ignored (the model is fit assuming that all circulating RNA are from asexual parasites). We would expect a higher proportion of circulating gametocytes in asymptomatic patients relative to symptomatic uncomplicated malaria (the transition to gametocyte production is delayed [5, 9]). The effect of gametocyte gene expression on the hpi estimation in this dataset has not been explored.

## 5 Conclusions

Persistence of infection is ubiquitous among human eukaryotic parasites, although the mechanisms vary. The evolutionary driver is simple: maximising transmission whilst evading host immune defences. It is often hypothesised that cytoadherence has evolved as a mechanism to avoid splenic clearance, so it is unclear how reducing cytoadherence would improve transmissibility, and therefore why it would be selected. The data reported in Andrade *et al* are interesting, but they might simply reflect a difference in synchronicity between acute and chronic malaria and the selection bias resulting from clinical presentation following synchronous schizogony. There is no need to postulate delayed cytoadherence beyond the known differences associated with fever [10].

### Supplementary information

The code used to generate Figures 1–3 is provided as an RMarkdown script hosted at the following github repository: https://github.com/jwatowatson/Andradereplication. The script downloads openly accessible data.

## Data Availability

All data and code are available online at: https://github.com/jwatowatson/Andrade_replication

https://journals.plos.org/plosbiology/article/file?type=supplementary&id=10.1371/journal.pbio.0000005.sd002

https://www.ncbi.nlm.nih.gov/geo/download/?acc=GSE148125&format=file&file=GSE148125%5FHS131%5Fnormalized%5Fcounts%2Etxt%2Egz

## Author contributions

All authors designed the paper by discussing the key concerns to be included. JAW analysed the data, wrote the simulations, and wrote the initial draft. All authors approved the final draft.

## Footnotes

[1 The computer sorts alphanumeric character strings in the same way as words are sorted in a dictionary. For example, when represented as character strings, the numbers 1, 19, 100 109 would be ranked in increasing order as 1, 100, 109, 19.

## References

[1] Andrade, C.M., Fleckenstein, H., Thomson-Luque, R., Doumbo, S., Lima, N.F., Anderson, C., Hibbert, J., Hopp, C.S., Tran, T.M., Li, S., et al.: Increased circulation time of Plasmodium falciparum underlies persistent asymptomatic infection in the dry season. Nature Medicine 26(12), 1929–1940 (2020)

[2] Bozdech, Z., Llinás, M., Pulliam, B.L., Wong, E.D., Zhu, J., DeRisi, J.L., Ward, G.: The transcriptome of the intraerythrocytic developmental cycle of Plasmodium falciparum. PLoS Biology 1(1), 5 (2003)

[3] Lemieux, J.E., Gomez-Escobar, N., Feller, A., Carret, C., Amambua-Ngwa, A., Pinches, R., Day, F., Kyes, S.A., Conway, D.J., Holmes, C.C., et al.: Statistical estimation of cell-cycle progression and lineage commitment in Plasmodium falciparum reveals a homogeneous pattern of transcription in ex vivo culture. Proceedings of the National Academy of Sciences 106(18), 7559–7564 (2009)

[4] Dondorp, A.M., Desakorn, V., Pongtavornpinyo, W., Sahassananda, D., Silamut, K., Chotivanich, K., Newton, P.N., Pitisuttithum, P., Smithyman, A., White, N.J., et al.: Estimation of the total parasite biomass in acute falciparum malaria from plasma PfHRP2. PLoS Medicine 2(8), 204 (2005)

[5] Fairley, H.N.: Sidelights on malaria in man obtained by subinoculation experiments. Transactions of the Royal Society of Tropical Medicine and Hygiene 40(5), 621–676 (1947)

[6] Wattanakul, T., Baker, M., Mohrle, J., McWhinney, B., Hoglund, R.M., McCarthy, J.S., Tarning, J.: Semimechanistic pharmacokinetic and pharmacodynamic modeling of piperaquine in a volunteer infection study with Plasmodium falciparum blood-stage malaria. Antimicrobial Agents and Chemotherapy 65(4), 01583–20 (2021). https://doi.org/10.1128/AAC.01583-20

[7] White, N., Chapman, D., Watt, G.: The effects of multiplication and synchronicity on the vascular distribution of parasites in falciparum malaria. Transactions of the Royal Society of Tropical Medicine and Hygiene 86(6), 590–597 (1992)

[8] Kwiatkowski, D.: Febrile temperatures can synchronize the growth of Plasmodium falciparum in vitro. The Journal of Experimental Medicine 169(1), 357–361 (1989)

[9] Bousema, J.T., Gouagna, L.C., Drakeley, C.J., Meutstege, A.M., Okech, B.A., Akim, I.N., Beier, J.C., Githure, J.I., Sauerwein, R.W.: Plasmodium falciparum gametocyte carriage in asymptomatic children in western Kenya. Malaria Journal 3(1), 1–6 (2004)

[10] Udomsangpetch, R., Pipitaporn, B., Silamut, K., Pinches, R., Kyes, S., Looareesuwan, S., Newbold, C., White, N.J.: Febrile temperatures induce cytoadherence of ring-stage Plasmodium falciparum-infected erythrocytes. Proceedings of the National Academy of Sciences 99(18), 11825–11829 (2002)

